# Street-food vendors and food safety: Behavioural predictors of safe vegetable washing in Accra, Ghana

**DOI:** 10.1101/2025.11.10.25339755

**Authors:** David Galibourg, Katherine V. Gough, Rebecca E. Scott, Jurgita Slekiene

**Affiliations:** Water Engineering and Development Centre (WEDC), School of Architecture, Building and Civil Engineering, Loughborough University, Loughborough, United Kingdom; International Water Management Institute, Accra, Ghana; Department of Geography and Environment, Loughborough University, Loughborough, United Kingdom; Department of Human Geography, Lund University, Lund, Sweden; Institute of Evolutionary Medicine, Anthropometrics & Historical Epidemiology Group, University of Zurich, Zurich, Switzerland

**Keywords:** Faecal contamination, food vendors, raw vegetables, potassium permanganate, behaviour change, motivation, social norms, RANAS, Accra, Ghana

## Abstract

**Background:** In Accra, Ghana, street food vendors soak leafy greens and other salad vegetables, eaten raw, in a basin of water to which they add salt, with the expectation that this will remove germs. While this method has long been the best available option, salt is less effective than other disinfectants. Potassium permanganate is one such disinfectant commonly used in French-speaking West Africa and other Low- and Middle-Income Countries, but its use to sanitise vegetables is virtually unknown in Ghana. This article aims to identify the factors that would induce street food vendors to use potassium permanganate instead of salt.

**Methods:** Using the Risk-Attitude-Norms-Abilities-Self regulation (RANAS) behaviour change approach, an exploratory sequential mixed-methods design consisting of formative interviews (N=20) and a quantitative survey of street food vendors (N=251) was conducted in 10 neighbourhoods of Accra to investigate which psychosocial factors best predict the adoption of potassium permanganate. An ordinal logistic regression analysis then determined the factors that are the most significant predictors of vendors’ commitment to using potassium permanganate.

**Results:** The interviews and survey showed that street food vendors are aware of the health risks associated with food contaminated with germs and prioritise hygiene. However, limited and sometimes inaccurate knowledge hinders their motivation to adopt safer practices. Our study revealed that personal norms, confidence in resuming the practice after an interruption, and others’ approval (injunctive norms) are significant predictors of street vendors’ commitment to using potassium permanganate. Our findings also suggest that education, training and financial (dis-)incentives are insufficient on their own to motivate the adoption of this safe practice.

**Conclusions:** Our results suggest that public authorities seeking to promote the use of potassium permanganate to sanitise vegetables eaten raw should integrate behaviour change strategies that leverage these predictors to enhance adoption. The article demonstrates the value of the RANAS approach for addressing food safety behaviours across urban vegetable value chains in low- and middle-income countries globally, aligning with WHO recommendations that emphasise socio-cultural dimensions alongside technical measures.

## Background

Consuming raw vegetables contaminated with faecal pathogens contributes meaningfully to 420,000 deaths and 600 million cases of illness, and the loss of 33 million Disability Adjusted Life Years (DALYs) globally due to foodborne diseases each year (1–3). The majority of this burden falls on Low- and Middle-Income Countries (LMIC), which lose USD100 billion annually due to inadequate food safety (4). In particular, contaminated lettuces, spring onions and other leafy vegetables are served raw by street food vendors, exposing their 2.5 billion daily consumers to a major public health risk (5–8).

In Accra, Ghana, consuming raw produce and street food poses a greater risk of exposure to faecal pathogens than drinking water or using public toilets (9). Yet, 85% of the population across all ages and income levels, including school-going children, consume such food almost daily (9–11). Vegetables accumulate pathogens as they transit from farm to fork. On the farm, pathogens are transferred from manure and irrigation water contaminated as a result of unsafe sanitation provision (12). Post-harvest, more pathogens are transferred due to traders’ and street food vendors’ poor personal hygiene and food handling practices, exacerbated by limited access to safe water and handwashing facilities at home and in markets (13). If farmers, traders and street food vendors had the capability, opportunity and motivation to adopt safe practices, combined these would reduce this contamination to a safe level (14–16). While most food safety interventions focus on vendors’ personal hygiene and safe handling practices to prevent post-harvest contamination, thorough washing of vegetables is necessary to reduce the pathogens transferred from the farm (17).

Guidance on how to wash vegetables to reduce contamination levels needs to be adapted according to the quality of the water accessible to food handlers. Where water quality is adequate, washing vegetables under running water is recommended and adding disinfectant is discouraged to avoid the risk of misuse (18). Where running water is unavailable or unsafe, it is recommended first to sanitise the water to be used in a container by adding a disinfectant (19). With the intermittent supply of piped water compromising its quality at the delivery point, only about 20% of Accra’s population relies on piped water for human consumption, with 75% buying sachets or bottled water, which are considerably more expensive (20,21). Consequently, street food vendors wash vegetables in three steps: first, they scrub them in a basin of piped water to remove sand and dirt, and any helminth eggs that stick to the leaves. Second, the vegetables are soaked in a basin of water to which salt (or vinegar, or lemon juice) is added with the intent to remove germs. Third, the vegetables are rinsed in a basin of plain piped water (22). As vendors believe salt is effective in killing germs and that adding the smallest amount is adequate (23), the dose of salt commonly used is insufficient to remove the germs present (24). Moreover, the water is often reused for several washing cycles, accumulating pathogens that can be re-transferred to vegetables washed later (20). Since most street food vendors favour the three-step wash method over washing under running water, and their use of salt is ineffective against germs, replacing salt with a more effective disinfectant offers a promising alternative (24).

Although behavioural frameworks are widely used in hygiene and sanitation research, their application to promoting the adoption of disinfectant for vegetable sanitation in informal food systems has received little attention. Chlorine is one of the disinfectants most commonly used globally to sanitise water and fresh vegetables (25). Potassium permanganate (KMnO_4_) is an alternative that is commonly used in French-speaking West Africa and other LMIC (26–29). Available as a powder (also called Condy’s crystals) or tablets (e.g., Permitabs), potassium permanganate can be purchased over the counter in pharmacies, as it is often used to treat skin fungal diseases. Soaking vegetables for between 5 and 10 minutes at room temperature in a pink to purple solution of 100 to 200 ppm (parts per million) of potassium permanganate reduces faecal coliform concentration by 0.6 to 3 log units (24) and 4.1 log units at a concentration of 1000 ppm (30). Potassium permanganate is particularly effective against Vibrio cholerae and reduces the concentration of some pesticide residues (31,32). Despite the effectiveness of using potassium permanganate, its adoption for washing vegetables largely depends on behavioural factors.

This article applies a theory-based behaviour change model to examine how the use of potassium permanganate can be promoted among street food vendors in Accra as a safer alternative to salt for washing raw vegetables. Specifically, it draws on the RANAS framework (Risk awareness, Attitudes, Norms, Abilities, and Self-regulation) (33), which offers a structured approach for assessing and analysing the psychosocial factors influencing behaviour in this context. The study addresses the following research questions: (1) What is the current status of street food vendors across the psychosocial factors defined by the RANAS model? (2) Which of these factors are associated with vendors’ commitment to using potassium permanganate instead of salt? (3) What are the implications of these findings for designing effective food safety interventions in informal market settings? Our findings are critical for public authorities aiming to improve food safety and protect public health by promoting effective vegetable sanitisation practices. By supporting and informing the design of targeted, evidence-based interventions, this article illustrates how safer practices can be made more attractive and sustainable in informal market settings. To our knowledge, this is one of only a few studies to apply a theory-driven behavioural framework to food safety, and the first to examine the adoption of potassium permanganate for vegetable sanitisation in informal urban food systems.

## Methods

### The RANAS model

The RANAS behavioural approach has been developed to focus on the psychosocial factors that influence behaviours relating to water, sanitation and hygiene in LMIC settings (33). These factors are derived from several psychological theories, including the Health Belief Model (34), Protection Motivation Theory (35), and Health Action Process Approach (36).

Drawing on Contzen and Mosler (37), Risk factors address vendors’ knowledge of health risks from food contaminated by faecal pathogens, perceptions of disease severity, and vulnerability. Attitudes include instrumental beliefs (e.g., costs and benefits of using potassium permanganate) and affective beliefs (e.g., emotional associations with the behaviour). Social Norms consist of descriptive norms (perceived peer behaviour), injunctive norms (perceived approval from peers and customers), and personal norms (personal obligation). Abilities refer to vendors’ confidence in performing the behaviour (confidence in performance, continuation, and recovery). Finally, Self-regulation refers to vendors’ intentions to perform the target behaviour (action planning) and maintain it (action control), along with their commitment to using potassium permanganate. Other contextual factors, such as the physical and institutional environments, are likely to influence behaviour adoption (e.g., endorsement by public authorities, product availability), but are only briefly mentioned here as they are beyond the scope of this behavioural analysis.

### Study design, sampling procedure and data collection

Following the RANAS approach, the study adopted an exploratory sequential mixed-method design informed by Creswell & Creswell (38) and Teddlie & Tashakkori (39). Creswell and Cresswell (38) advocate that exploratory sequential designs are particularly valuable when existing instruments may not be culturally appropriate or when local contextual factors need to be understood before quantitative measurement. Moreover, as Teddlie and Tashakkori (39) emphasise, the qualitative phase should generate contextually relevant constructs that inform the quantitative phase. Such an approach was adopted to adapt the RANAS model to the specific context of Ghanaian street food vendors. Qualitative semi-structured interviews were used to explore the perceptions of a small sample of street food vendors (n=23) regarding hygiene, food safety, and vegetable washing methods. These data were then analysed to inform the design of a contextually sensitive structured questionnaire. The questionnaire was administered to a larger sample of street food vendors (n=251) to quantitatively measure the strongest predictors of their commitment to use potassium permanganate.

The interview guide consisted of 24 questions. The first set of questions covered socio-demographics, current practices and knowledge, and disinfectant preferences, aiming to provide context for understanding respondents’ answers to the later questions, which explored the factors included in the RANAS model. The interview guide was piloted with a few vendors to identify any issues with comprehension or length. Based on the pilot feedback, the guide was shortened to accommodate vendors’ limited time for interviews.

After securing authorisation from the relevant authorities, three interviews were conducted in April 2023 with vendors operating in three schools selected for this study by the School Health Education Programme in the Accra Metropolitan Assembly. These were Tesano Police Depot Primary School, Mamprobi Socco H/H Basic School, and Derby Avenue Roman Catholic Basic School. Another 20 interviews were conducted with vendors in February 2024, following a targeted purposive-convenience sampling approach across four neighbourhoods in Accra. Low and middle-income areas were selected in each neighbourhood to represent the broader segment of the population: Mataheko and Osu, based on a prior study highlighting the significance of street food contamination in these neighbourhoods (10,40), and Madina and Nima for their popularity among salad vendors. Five vendors were selected for interview in each neighbourhood based on their sale of salads as part of their food offerings, typically served with rice dishes, or on their own. Gender was not a specific criterion, as over 90% of street food vendors are women; however, male vendors were included if available.

Where possible, the interview was conducted with the person responsible for washing vegetables, either the vendor themselves or one of their employees. Interviewees were informed of the study’s objectives and permission to audio record was requested. Most vendors discussed openly in front of their employees and customers (if any were present during the interview). Under the direction of the lead author, the interviews were conducted with the support of two trained local research assistants, in the respondent’s preferred language. Live interpretation by the research assistants enabled the lead author to take fieldnotes and adapt the interview in real-time. The interviews lasted 20-50 minutes, depending on vendor availability and interest. The audio recordings were subsequently transcribed and translated into English. The transcripts and fieldnotes were then coded and analysed by the lead author using the RANAS model as an analytical framework. This facilitated the qualitative characterisation of vendors’ current knowledge and practices and informed the design of a quantitative questionnaire.

Following pre-testing, the quantitative questionnaire survey was conducted in February and March, 2024 with street food vendors operating in a broader range of neighbourhoods across the Accra metropolis: Abeka-Lapaz (n=25), Kole (n=25), Kotobabi (n=25), Labadi (n=27), Maamobi (n=25), Madina (n=25), Mataheko-Kaneshie (n=25), New Town (n=25), Nima (n=25), and Osu (n=24) (see Figure 1). In addition to the four locations where interviews were conducted, other areas were included to ensure a broad coverage of the study region, stretching from the city centre to the outskirts along both north-south and east-west axes, encompassing areas with different population densities and diverse zoning. A 2016 survey identified 674 street food vendors in areas overlapping with our study sites (41). Given that an estimated 35% of vendors serve rice-based dishes (42,43, quoted in 44), approximately 236 vendors in these areas would be relevant to our study. Our sample of 251 respondents represents comprehensive coverage of vendors serving rice-based dishes topped with salad in these areas, suggesting either growth in this vendor segment since 2016 or a slightly broader geographic coverage in our sampling approach. Vendors included in the interviews were excluded from the survey sampling to avoid potential priming effects and reduce bias.

**Figure 1.**
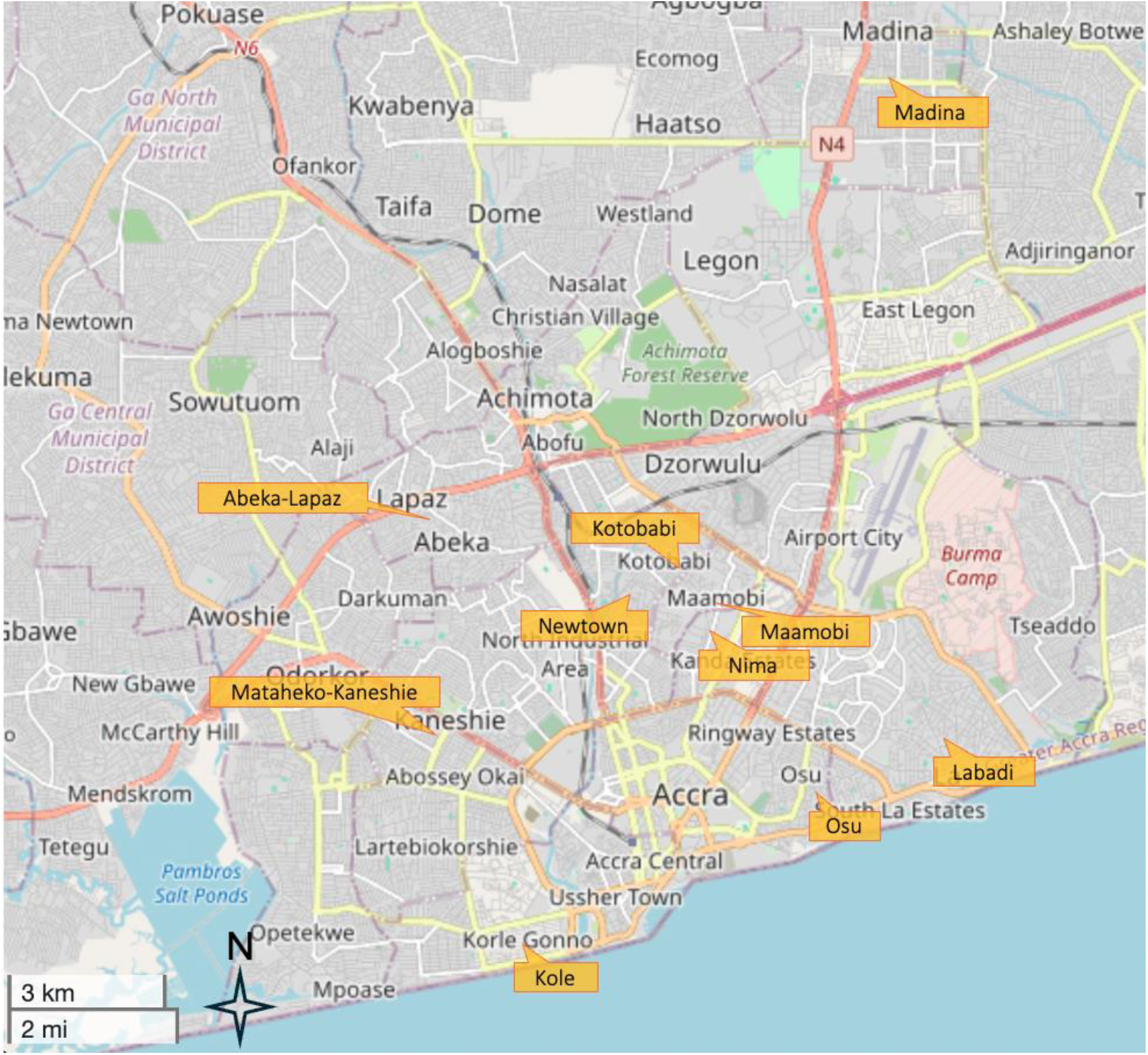
Locations of the street food vendors surveyed (Map data source: OpenStreetMap)

The aim of the quantitative questionnaire was to explore psychosocial factors included in the RANAS model to assess their influence on vendors’ willingness to use potassium permanganate for washing raw vegetables. Each factor was assessed using one or more dichotomous or Likert-type questions. As vendors currently use salt for washing vegetables, the survey measured their commitment to switch to potassium permanganate. It was assessed using a self-reported 3-point Likert scale (1: not committed at all, 2: a little committed, 3: very committed), which was used as the outcome variable.

The statistical analyses were conducted using IBM SPSS 29 Statistics software (45) and R version 4.5.1 (46). Given the ordinal nature of the outcome variable (3-point Likert scale), an ordinal logistic regression was conducted as the primary analysis, treating commitment as ordered categories (1 < 2 < 3). Prior to the multivariable modelling, preliminary diagnostics were performed to assess construct overlap (Spearman correlations), multicollinearity (VIF), and univariate associations (Kruskal-Wallis tests). Sensitivity analyses using binary logistic regression (two coding schemes) and linear regression validated the robustness of findings. Ordinal logistic regression was selected as the primary approach given its theoretical appropriateness for ordered categorical outcomes (47). This approach preserves the ordinal structure of the data without imposing assumptions of equal intervals between response categories. A complete case analysis was used (n=251). The detailed diagnostic results and method comparisons are provided in Tables 7-11 in the Supplementary Material.

Four contextual factors (age, education, willingness to pay, and hygiene knowledge) were also explored. Here, too, gender was not explicitly examined, as almost all the vendors were female (n=233). Respondents’ age was categorised into five ordinal groups, ranging from 1 (less than 20 years) to 5 (50 years or older). Education was recorded on a 9-point ordinal scale from 1 (no formal education) to 9 (university level). Willingness to pay was assessed using a 5-point scale ranging from 1 (GHS^1^ 0.50) to 5 (more than GHS 5). Hygiene knowledge was assessed through multiple-choice questions. Dichotomous (yes/no) items captured whether respondents used running or stored water, added salt or vinegar to wash vegetables, and knew about chlorine and potassium permanganate. The frequency of handwashing with water and soap, as well as washing vegetables with salt or vinegar, was measured on a 5-point Likert scale from 1 (almost never) to 5 (almost always). Psychosocial factors were measured using one or more items, using bipolar Likert scales, either 3- or 5-point, depending on the construct (see Error! Not a valid bookmark self-reference.).

Data were collected by nine enumerators fluent in local languages, supervised by the lead author. The survey, initially developed in English, was translated into local languages and adjusted to be culturally appropriate in collaboration with the team during two days of intensive training. The training programme covered the study’s goals and key concepts, and the theoretical background, principles and constructs of the RANAS method. The survey was administered using the mobile application KoboToolBox (48). In each neighbourhood, enumerators approached vendors, assessed their eligibility, and obtained informed consent before collecting the demographic and behavioural data.

The study adhered to ethical protocols approved by Loughborough University’s ethical review committee and the Institutional Review Board of the International Water Management Institute (IWMI) in Ghana. All team members completed UNICEF’s “Introduction to Ethics in Evidence Generation” programme. The lead author monitored data quality, regularly providing and collecting feedback. In the cases of three respondents, missing data for one question were handled using pairwise deletion, allowing the remaining variables to be included in the analyses.

## Results and discussion

### Socio-demographics

Street food vending in Ghana is predominantly a female activity, reflecting the influence of traditional gender norms and expectations (49). Consequently, the majority (92%) of the 251 street food vendors who participated in this study were female, with the most common age range (41%) being 30-39 (see Table 2). Most vendors received some formal education, especially among the younger age categories, with 81% reaching Junior High School (JHS) level or higher. Compared to a decade ago, street food vendors appear to be more educated: the proportion of vendors without formal education has decreased by two-thirds, while those with Senior High School (SHS) or tertiary education have doubled (41). Despite this trend, formal vocational training remains limited, with only 11% of vendors having received such training, though this is twice the figure reported a decade ago. Most respondents chose to take the survey in Twi (62%).

**Table 1.** Psychosocial factors considered in the RANAS approach.

| RANAS factors | Example questions | Scale |
| --- | --- | --- |
| Risk factors: individuals' understanding and awareness of the health risk |  |  |
| Factual knowledge | Can germs come from irrigation water? | From 0 (no correct answer) to 9 (all answers correct) |
| Perceived vulnerability | How often do you think you or other adults can get sick after eating a salad that has germs? | From 1 (almost never) to 4 (very often) |
| Perceived severity | How sick do you think you or other adults can be from eating a salad that has germs? | From 1 (not sick at all) to 5 (very sick) |
| Attitude factors: individuals' positive or negative stance towards a behaviour |  |  |
| Beliefs about food safety | What is more important to you about your salads: quantity, safety, or tastiness? Scale applied to each option | From 0 (quantity or taste first) to 1 (safety first) |
| Instrumental beliefs | Do you think using potassium permanganate instead of salt could be a problem if it is a little more expensive? | From 1 (not a problem) to 3 (a big problem) |
| Affective beliefs (feelings) | If you had to start washing salad vegetables with potassium permanganate, how would that make you feel? | From 1 (very worried) to 5 (very comfortable) |
| Willingness to pay | How much would you be willing to pay per day for potassium permanganate? | From 1 (0.5 cedi) to 5 (>5 cedis) |
| Norm factors: individuals' perception of the social pressure towards a behaviour |  |  |
| Descriptive norm (others' behaviour) | How many other street food vendors do you think would be willing to use potassium permanganate if it makes salads safer? | From 1 (no-one) to 4 (almost all) |
| Injunctive norm (others' (dis-)approval) | People who are important to you, like family members, or religious leaders, to what extent would they approve that you wash your salad vegetables with potassium permanganate? | From 1 (they would not approve) to 3 (they would approve) |
| Personal norm (personal importance) | To you personally, how important is it to make salad safer by washing vegetables with potassium permanganate? | From 1 (not important at all) to 3 (very important) |

Table 1 Psychosocial factors considered in the RANAS approach (continued)
| RANAS factors | Example questions | Scale |
| --- | --- | --- |
| Ability factors: individuals' confidence in their ability to practise a behaviour |  |  |
| Confidence in performance | How confident are you that you would be able to measure 5 litres of water? | From 1 (not confident) to 3 (very confident) |
| Confidence in continuation | How confident are you that you would be able to use potassium permanganate continuously? | From 1 (not confident) to 3 (very confident) |
| Confidence in recovery | If you had to stop using potassium permanganate because it was unavailable at the pharmacy, how confident are you that you would start using it again? | From 1 (not confident) to 3 (very confident) |
| Self-regulation factors: individuals' attempts to plan and self-monitor a behaviour and overcome barriers |  |  |
| Action planning | How do you plan to use potassium permanganate? | From 0 (can describe) to 1 (cannot describe) |
| Action control | How difficult or easy do you think it would be to always use the correct quantity of potassium permanganate and water and the correct time when washing salad vegetables? | From 1 (very difficult) to 4 (very easy) |
| Commitment | If the authorities you trust told you that washing vegetables with potassium permanganate makes salads very safe, what is the likelihood of you continuing to use it? | From 1 (not committed at all) to 3 (very committed) |

**Table 2.** Socio-demographic characteristics of respondents.

| Characteristics | n | % |
| --- | --- | --- |
| **Gender** |  |  |
| Female | 233 | 92.4 |
| Male | 19 | 7.6 |
| **Age (years)** |  |  |
| <20 | 5 | 2.0 |
| 20-29 | 70 | 27.9 |
| 30-39 | 103 | 41.0 |
| 40-49 | 58 | 23.1 |
| 50+ | 16 | 6.0 |
| **Education level** |  |  |
| None | 13 | 5.2 |
| Can read but not write | 1 | 0.4 |
| Can read and write | 1 | 0.4 |
| Koranic | 9 | 3.6 |
| Primary | 25 | 9.6 |
| Junior High School (JHS) | 69 | 27.5 |
| Vocational | 27 | 10.8 |
| Senior High School (SHS) | 90 | 35.9 |
| Tertiary/university | 17 | 6.8 |
| **Language chosen for the questionnaire** |  |  |
| Twi | 157 | 62.5 |
| English | 69 | 27.5 |
| Ga | 25 | 10.0 |

Note. N=251.

### Self-reported behaviours

Although 57% of street food vendors reported ‘almost always’ having access to water and soap for handwashing, 43% encountered occasional or frequent difficulties. This echoes observations that hygiene and food safety infrastructure are often inadequate in markets (50,51). Formative interviews confirmed that while public toilets are available and their operators ensured a water supply, soap was often missing.

As seen in Table 3, most vendors (94%) reported washing vegetables in a basin, typically adding salt (91%) rather than using running water (6%), while about half use vinegar occasionally (53%). Over half (55%) of vendors reported ‘almost never’ washing salad vegetables without adding salt or vinegar, and 45% admitted to doing this at least sometimes, through a lack of planning or to save costs. Interviewees noted that vinegar, often considered a premium disinfectant, was mainly reserved for family meals, esteemed guests, or special occasions, as also observed by Rheinlander (23). This highlights the gap between vendors’ daily practices and public authorities’ recommendations, which advocate washing salad vegetables under running water (52), a practice constrained by both cost and infrastructure limitations. Reusing the same water in a basin for washing multiple times raises significant risks of cross-contamination.

**Table 3.** Self-reported behavioural frequencies and risk knowledge.

| Question | n | % |
| --- | --- | --- |
| Do you wash your salad vegetables in a basin or under running water? |  |  |
| Basin | 235 | 93.6 |
| Running water | 16 | 6.4 |
| Do you add salt to the water when you wash your salad vegetables in a basin? |  |  |
| No | 23 | 9.2 |
| Yes | 229 | 90.8 |
| Do you add vinegar to the water when you wash your salad vegetables in a basin? |  |  |
| No | 119 | 47.4 |
| Yes | 133 | 52.6 |
| Have you ever washed salad vegetables in a basin without salt or vinegar? |  |  |
| Almost never | 138 | 55.0 |
| Sometimes | 89 | 35.5 |
| Often | 13 | 5.2 |
| Almost always | 11 | 4.4 |

Note. N=251.

### Risk factors

Street food vendors exhibited greater awareness of contamination risks linked to the market environment, such as exposure to dust, flies, and handlers’ dirty hands, compared to risks originating on the farm, such as contamination from soil or irrigation water. A vendor in her 40s, with JHS education, commented that she was “Not sure about the sickness caused by germs”, while another, in her 20s, added, “It is solely the chemicals [chemical residue] that bring [diseases] on vegetables”. A vendor in her 30s reported using salt to remove sand and germs, which she described as maggots and lice sometimes found on vegetables. This aligns with other studies, which indicated that less than 20% of Accra’s street food vendors associate diarrhoea with faecal pathogens (53), believing that they are not harmful (54). The majority of respondents think that consuming contaminated salads can lead to one or more common mild to moderate health issues, such as diarrhoea (28%), stomachache (29%), or vomiting (30%). Most respondents acknowledge that contaminated salads can transmit cholera (83%) or typhoid fever (51%), two diseases that require medical attention. However, only 10% believe contaminated salads pose health risks serious enough to warrant consulting a health specialist. Vendors may perceive food-borne disease severity as low because their experience with cholera is limited to outbreaks distant in time and location. Diarrhoea is often perceived as a routine discomfort rather than a serious illness, making it harder to link to consuming contaminated salads. The vast majority of vendors (95%) think children are more likely to contract diarrhoeal diseases (i.e., vulnerability) and that these could be more serious (i.e., severity) than in adults (90%). Many vendors believe individuals develop immunity to germs, whereas a more accurate perspective is that adults are survivors of diarrhoeal diseases, which remain a leading cause of death among children under five in Ghana and other LMIC (55,56).

Vendors’ understanding of food contamination risks is fragmented and sometimes inaccurate. Some believe that contamination can be introduced in the market but not on the farm, that unwashed vegetables can cause not only diarrhoea but also malaria, and/or that the smallest amount of salt is enough to remove anything harmful from vegetables. Vendors are aware of the health risk of contaminated food, which they believe can be common but generally not severe. This can lead them to believe that personal hygiene and adequate handling are sufficient to ensure food safety, as these actions prevent pathogens from being introduced at the market level. However, only careful washing of vegetables can reduce the concentration of pathogens transferred from the farm. Careful washing is a critical control point as food safety status does not necessarily align with the visible neatness of the vendor, their booth and surroundings or the freshness of the vegetables, which vendors and customers rely on as sensory proxies of food hygiene and quality (57).

### Vendors’ preferences regarding disinfectant options

Street food vendors were shown two disinfectants purchased from local pharmacies: an effervescent chlorine-based tablet (Aquatabs) in its blister pack (58), and potassium permanganate powder in a small vial. They were informed that Aquatabs and household bleach both rely on chlorine compounds to kill germs. It was also explained that chlorine and potassium permanganate are more effective than salt for sanitising vegetables. Approximately 44% of respondents were familiar with Aquatabs, while only 15% knew about potassium permanganate. When asked which disinfectant they would prefer to use (between Aquatabs tablets, potassium permanganate and liquid household bleach, commonly referred to as Parazone, a popular brand name in Ghana) instead of salt, most respondents (77%) declared their preferred option would be Aquatabs (Table 4). Potassium permanganate was the second choice for 65% of respondents. Using liquid bleach as a food disinfectant was overwhelmingly rejected by 92%.

**Table 4.**
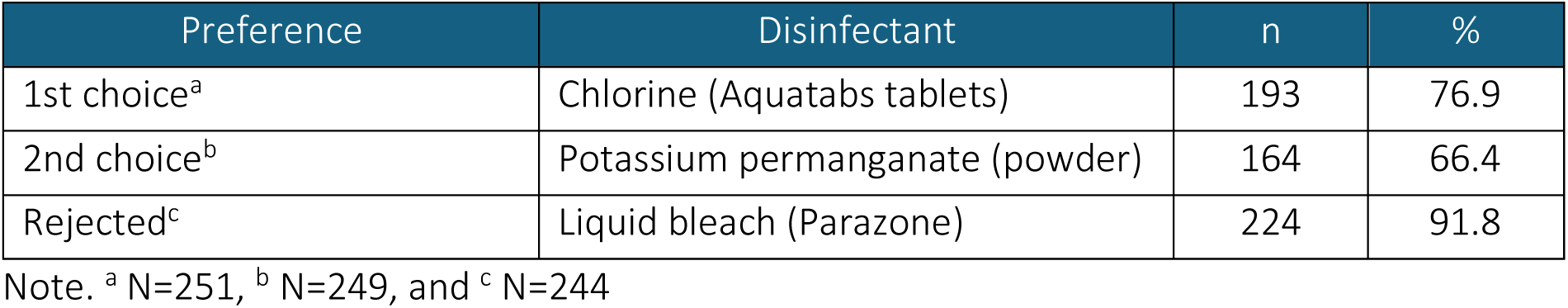
Preferences regarding disinfectants to sanitise vegetables instead of salt.

| Preference | Disinfectant | n | % |
| --- | --- | --- | --- |
| 1st choice<sup>a</sup> | Chlorine (Aquatabs tablets) | 193 | 76.9 |
| 2nd choice<sup>b</sup> | Potassium permanganate (powder) | 164 | 66.4 |
| Rejected<sup>c</sup> | Liquid bleach (Parazone) | 224 | 91.8 |

Note. <sup>a</sup> N=251, <sup>b</sup> N=249, and <sup>c</sup> N=244

This preference for salt over any chemical disinfectant is reflected in the comment made by a vendor from New Town, who was in her 20s and studied to the JHS level: “[I] only believe in using the Ada salt [coarse salt harvested in the Ada area]”. A vendor from Kole, in her 30s, also with JHS education, added: “These products [chlorine or potassium permanganate] will kill us all. Salt is healthier since [I’m] still alive”. Vendors view salt as a natural, ‘healthy’ disinfectant, akin to vinegar, which they consider effective but more expensive. Some respondents indicated being taught at vocational school how to wash vegetables with vinegar if possible and, if not, with salt. Many vendors stated that their mothers and grandmothers taught them to use salt for washing vegetables. Whether it is salt, vinegar or lemon juice, respondents believe that only edible disinfectants should be used with food. As a respondent in her 20s with JHS education explained: “Even though I said I will use both Aquatabs and potassium permanganate, I am a bit resistant because I thought it was poison.”

The preference for Aquatabs is likely due to respondents’ familiarity with the product, which the government occasionally distributes during water supply disruptions caused by flooding. The convenient blister-packaging, unlike loose powder, may have influenced preference by making Aquatabs “Look like a medicine,” as noted by a female in her 30s with SHS education. However, the smell of chlorine is stronger when Aquatabs are used to sanitise vegetables, as the manufacturer recommends higher doses for vegetable sanitation than for water purification. Interviewees consistently rejected bleach (or any products that smell of chlorine) to sanitise vegetables as they associate it with cleaning toilets. As one vendor from Nima (in her 20s with no formal education) put it, “I don’t like that Aquatabs smell like Parazone.” Given the strong association of the chlorine smell with toilet cleaning, vendors are likely to reject Aquatabs or other chlorine-based products at the concentrations needed to sanitise vegetables. Potassium permanganate is, however, odourless and users who were interviewed reported having no concern about its purple colouration when sanitising vegetables, as the colour does not transfer to the vegetables.

### Attitude factors

Most respondents prioritised food safety (74%) over its palatability (i.e., tasty) or quantity (i.e., plenty). Vendors and consumers often conflate “food safety”, the terminology used for the survey, with food, vendor and stall neatness, as indicated earlier. Taste is important to satisfy customers, who often prioritise it over quantity. In Accra, vendors match quantity and price as customers specify the amount of each ingredient they want, such as requesting GHS 10 worth of rice and GHS 2 worth of salad.

There was no salient trend as to respondents feeling either worried or comfortable with the idea of using potassium permanganate. A respondent in her 20s, with SHS education, reported the often-expressed view that she “spent most of her life using salt” and feels “it will be difficult to make a change”. Others commented that they would be more at ease using potassium permanganate if more education were provided. Yet, in line with prioritising food safety, 86% of vendors think using potassium permanganate would be at least a little advantageous if it is more effective than salt. Customer and peer acceptance (62%) and the accessibility of potassium permanganate (55%) are perceived as greater barriers than cost (38%) (Table 5). Some female interviewees recalled potassium permanganate being used as a childhood treatment for fungal genital infections administered by their mothers, though its decreasing availability in pharmacies was indicated. While the reasons are unclear, studies have documented cases of its misuse leading to severe genital burns, in the mistaken belief that it induces abortion (59,60). Out of 20 pharmacies visited in February 2024 in Madina, Mataheko, Nima and Osu, only six sold Aquatabs and three supplied potassium permanganate. Ensuring accessibility to potassium permanganate would be key to promoting its use among street food vendors.

**Table 5.**
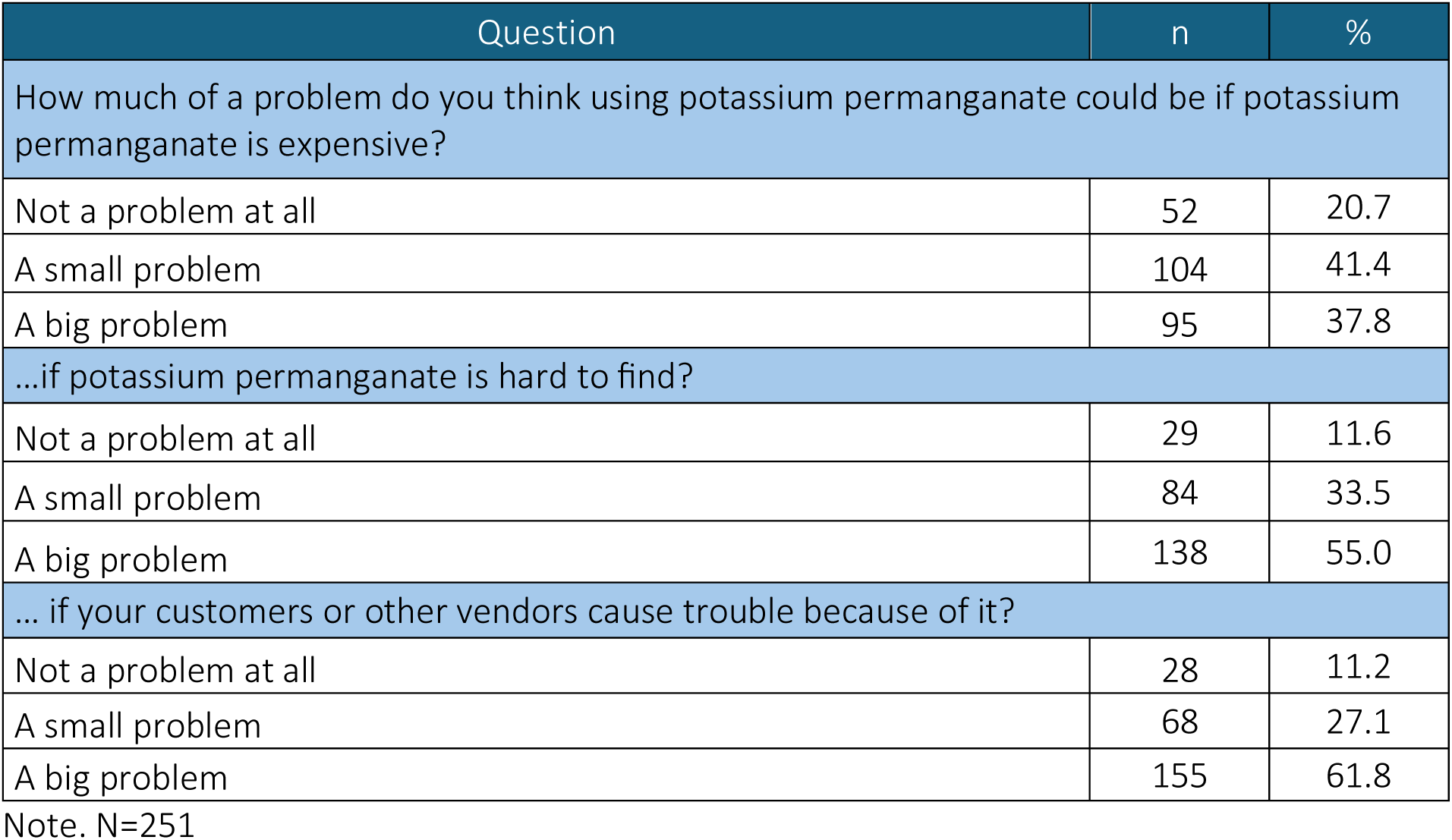
Attitude towards the use of potassium permanganate.

About two-thirds of respondents would be willing to pay more than 2.5 cedis per day on potassium permanganate. A vendor in her 20s operating in Mataheko commented: “If permanganate is cheap, like about 10 to 30 cedis, I’ll be willing to use it.” When asked about their willingness to pay, some respondents commented that they would pass the cost along to their customers. To put vendors’ willingness to pay for potassium permanganate into perspective, a rice dish with salad costs GHS 10-15. While the currency is volatile, a 2016 study estimated that vendors spend around GHS 100 on ingredients daily and generate sales of approximately GHS 160 (41).

### Norm factors

Over 90% of respondents declared it would be personally important to make salads safer by using potassium permanganate (43% a little important and 48% very important). Moreover, about 43% think that people important to them would approve of their use of potassium permanganate to make salads safer. Yet, 71% assume that few, if any, of their peers share similar views. Salt use is a deeply ingrained tradition, and more effective ‘chemical’ disinfectants are met with scepticism. Even those familiar with potassium permanganate have never heard of its use for sanitising vegetables. This sentiment is reflected in the words of a respondent in her 40s, without formal education: “My grandma taught [me] that salt is the only way to keep salad vegetables clean and that potassium permanganate should not be eaten but used for external purposes.” Tradition and conformism are strong values in Ghanaian society (61). This could explain why vendors are privately open to using potassium permanganate, especially if following official advice, but believe that most of their peers would not, despite many sharing the same willingness.

### Ability factors

Vendors were first told how to use potassium permanganate instead of salt during the second wash, i.e., dissolving a tip-of-a-knife amount of potassium permanganate in 5 litres of clean water (e.g., piped water) and soaking vegetables for 10 minutes. Following this, over 90% of respondents felt at least somewhat confident in their ability to use potassium permanganate correctly. To ensure correct dilution, vendors were advised to use a 2.5 L empty soda bottle to measure 5 L. Interestingly, while the amount of salt that vendors report using can be approximate, a respondent in her 40s, without formal education, suggested a small plastic measuring spoon should be provided with potassium permanganate, as ‘tip of a knife’ is not a precise measurement. Another vendor in her 20s, without formal education, indicated she might not have the time to let vegetables soak for 10 minutes. Most respondents expressed some level of confidence in their ability to continue using potassium permanganate, with 45% reporting they were “A little confident”. Similarly, 46% felt “A little confident” and 32% were “Very confident” about resuming their use after a temporary shortage.

### Self-regulation factors

A quarter of respondents stated that they would ensure they purchased additional potassium permanganate before running out, whereas 57% admitted they would revert to using salt in such instances. Most respondents felt it would be either easy (41%) or very easy (20%) to use potassium permanganate correctly. Some respondents, including one with vocational training, noted that they would feel more confident after personally trying out potassium permanganate. In terms of commitment, 43% reported they would be a little committed, and 47% very committed to using potassium permanganate instead of salt, and the remainder were not committed at all. Some respondents said their commitment would increase if they could first test potassium permanganate, especially if endorsed by the Food and Drug Authority (FDA) and with proper education regarding its use. These comments highlight vendors’ need for clear guidance from trusted sources, which contrasts with consumers’ lack of trust in the competence of public institutions to ensure food safety (62). Vendors’ reluctance to switch to potassium permanganate also reflects familiarity bias and a tendency to avoid uncertainty. For instance, a respondent in her 40s, with vocational training, stated that she was “More interested in Aquatabs” because she “had heard of it before”. Another one, in her 40s, with JHS education, was “Very sceptical to try it [potassium permanganate]” because she is “[only] now hearing of it”.

### Modelling street food vendors’ adoption of potassium permanganate

The weight of the different RANAS factors was determined through an ordinal logistic regression analysis, with the respondents’ commitment to using potassium permanganate as the dependent variable (outcome) and 16 RANAS psychosocial factors as independent variables (predictors) to explore which psychosocial factors determine safe vegetable washing practices among street food vendors (see Error! Reference source not found.).

Correlation analyses confirmed that predictors measured distinct constructs from the outcome. Personal norms showed the strongest association with commitment (rs = 0.62), followed by injunctive norms (rs = 0.57) and maintenance confidence (rs = 0.53). While the correlation between personal norms and commitment is substantial, these constructs are theoretically distinct within the RANAS framework: personal norms reflect internalised values about the importance of a behaviour. In contrast, commitment represents behavioural intention to act. This relationship, where perceived importance predicts behavioural intention, is consistent with established health behaviour change theories (36,63) and represents an expected theoretical pathway rather than measurement redundancy. Moreover, the correlation remained below the threshold for construct overlap concerns (r < 0.7), and VIF values confirmed acceptable multicollinearity (all VIFs < 5, with personal norms VIF = 2.74).

The model demonstrated strong explanatory power: the likelihood ratio test comparing the full model to the null model was highly significant (χ² = 179.69, df = 21, p < .001). McFadden’s pseudo-R² of 0.37 indicated excellent model fit (values of 0.2-0.4 are considered excellent for ordinal regression), and the Nagelkerke pseudo-R² of 0.60 demonstrated that the model explained 60% of the variance in commitment. The model achieved 75% prediction accuracy, representing a substantial improvement over the baseline (54% reduction in classification error).

The ordinal logistic regression identified four significant predictors of commitment (Table 6). ‘Personal norms’ was the strongest predictor (β = 1.32, OR = 3.73, 95% CI [1.80, 7.73], p < .001), indicating that vendors who viewed using potassium permanganate as personally important had 3.7 times higher odds of greater commitment. ‘Confidence in recovery’ was also significant (β = 0.92, OR = 2.51, 95% CI [1.40, 4.51], p = .002), suggesting vendors who are confident in their ability to resume use after interruptions demonstrated 2.5 times higher commitment odds. ‘Injunctive norms’ (β = 0.60, OR = 1.82, 95% CI [1.03, 3.21], p = .039) indicated that perceived social approval from influential figures, such as family members or religious leaders, positively influenced commitment. Finally, beliefs about the difficulty of finding potassium permanganate (β = -0.56, OR = 0.57, 95% CI [0.33, 0.97], p = .038) showed that concerns about accessibility reduced commitment odds by 43%.

**Table 6.**
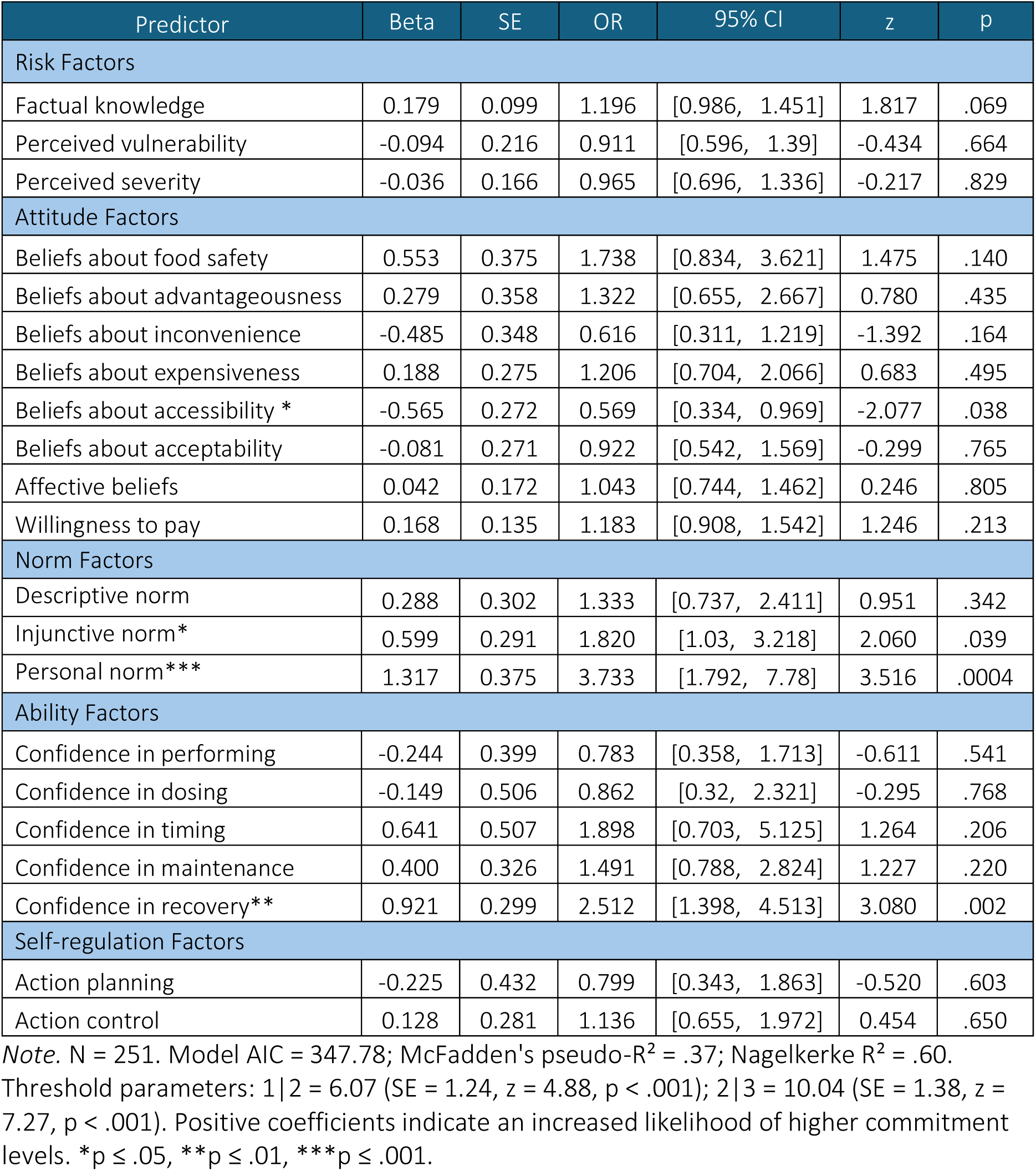
Ordinal logistic regression predicting commitment to using potassium permanganate.

| Predictor | Beta | SE | OR | 95% CI | z | p |
| --- | --- | --- | --- | --- | --- | --- |
| **Risk Factors** |  |  |  |  |  |  |
| Factual knowledge | 0.179 | 0.099 | 1.196 | [0.986, 1.451] | 1.817 | .069 |
| Perceived vulnerability | -0.094 | 0.216 | 0.911 | [0.596, 1.39] | -0.434 | .664 |
| Perceived severity | -0.036 | 0.166 | 0.965 | [0.696, 1.336] | -0.217 | .829 |
| **Attitude Factors** |  |  |  |  |  |  |
| Beliefs about food safety | 0.553 | 0.375 | 1.738 | [0.834, 3.621] | 1.475 | .140 |
| Beliefs about advantageousness | 0.279 | 0.358 | 1.322 | [0.655, 2.667] | 0.780 | .435 |
| Beliefs about inconvenience | -0.485 | 0.348 | 0.616 | [0.311, 1.219] | -1.392 | .164 |
| Beliefs about expensiveness | 0.188 | 0.275 | 1.206 | [0.704, 2.066] | 0.683 | .495 |
| Beliefs about accessibility * | -0.565 | 0.272 | 0.569 | [0.334, 0.969] | -2.077 | .038 |
| Beliefs about acceptability | -0.081 | 0.271 | 0.922 | [0.542, 1.569] | -0.299 | .765 |
| Affective beliefs | 0.042 | 0.172 | 1.043 | [0.744, 1.462] | 0.246 | .805 |
| Willingness to pay | 0.168 | 0.135 | 1.183 | [0.908, 1.542] | 1.246 | .213 |
| **Norm Factors** |  |  |  |  |  |  |
| Descriptive norm | 0.288 | 0.302 | 1.333 | [0.737, 2.411] | 0.951 | .342 |
| Injunctive norm* | 0.599 | 0.291 | 1.820 | [1.03, 3.218] | 2.060 | .039 |
| Personal norm*** | 1.317 | 0.375 | 3.733 | [1.792, 7.78] | 3.516 | .0004 |
| **Ability Factors** |  |  |  |  |  |  |
| Confidence in performing | -0.244 | 0.399 | 0.783 | [0.358, 1.713] | -0.611 | .541 |
| Confidence in dosing | -0.149 | 0.506 | 0.862 | [0.32, 2.321] | -0.295 | .768 |
| Confidence in timing | 0.641 | 0.507 | 1.898 | [0.703, 5.125] | 1.264 | .206 |
| Confidence in maintenance | 0.400 | 0.326 | 1.491 | [0.788, 2.824] | 1.227 | .220 |
| Confidence in recovery** | 0.921 | 0.299 | 2.512 | [1.398, 4.513] | 3.080 | .002 |
| **Self-regulation Factors** |  |  |  |  |  |  |
| Action planning | -0.225 | 0.432 | 0.799 | [0.343, 1.863] | -0.520 | .603 |
| Action control | 0.128 | 0.281 | 1.136 | [0.655, 1.972] | 0.454 | .650 |

Note. N = 251. Model AIC = 347.78; McFadden's pseudo- $R^2 = .37$ ; Nagelkerke $R^2 = .60$ .
Threshold parameters: 1 | 2 = 6.07 (SE = 1.24, $z = 4.88$ , $p < .001$ ); 2 | 3 = 10.04 (SE = 1.38, $z = 7.27$ , $p < .001$ ). Positive coefficients indicate an increased likelihood of higher commitment levels. \* $p \leq .05$ , \*\* $p \leq .01$ , \*\*\* $p \leq .001$ .

Sensitivity analyses using linear regression and binary logistic regression (with two coding schemes) tested whether findings were robust to different modelling assumptions. ‘Confidence in recovery’ remained significant across all four analytical approaches. ‘Personal norms’ was significant in ordinal regression, linear regression, and binary logistic regression with 1 vs 2-3 coding, though not significant in the 1-2 vs 3 coding (p = .118). This convergence of the two strongest predictors across multiple methods strengthens confidence that these represent genuine relationships rather than artefacts of analytical choice (see Table S8 in Supplementary Material for detailed comparison).

### Implications

Four factors significantly predicted vendors’ commitment: personal norms (OR = 3.73), confidence in recovery (OR = 2.51), injunctive norms (OR = 1.82), and beliefs about accessibility (OR = 0.57, indicating concerns about availability reduced commitment odds by 43%). These predictors are more influential than aspects traditionally emphasised in food safety interventions. Traditional targets showed weaker, non-significant effects: factual knowledge (OR = 1.20, p = .069), confidence in performance (ORs = 0.78–1.90, all p > .20), and willingness to pay (OR = 1.18, p = .213). Consequently, while education, training, and financial support are likely necessary, the RANAS model suggests they are insufficient on their own to drive sustained adoption of potassium permanganate.

We conducted post-hoc mediation analyses to explore whether emotional responses may enhance the salience and internalization of normative messages, as suggested by Packard and Schultz (64). Our analyses show that personal and injunctive norms fully mediate the relationship between emotional responses about food safety and commitment (see Tables S9 and S10 and Figure S1 in Supplementary Material). If further research was to confirm a causal relationship, this would suggest that interventions incorporating emotional appeals, such as pride in protecting customers, or concern about foodborne illness, might activate normative beliefs more effectively than purely information-based approaches.

Building on this understanding, practical strategies could include fostering a positive group identity and informing vendors that their peers and trusted authorities (e.g., representatives of the Food and Drug Authority, vendor association, or public health agents) or respected champions (e.g., chefs whom vendors follow on social media) approve potassium permanganate use. Additionally, ensuring potassium permanganate is readily available and accessible in local pharmacies and markets is essential to address practical barriers identified by vendors. This also highlights a key challenge: interventions that attempt to address all factors risk diluting their message, while those focusing on a single factor may not generate enough momentum for meaningful behaviour change.

### Limitations

This study has several limitations that should be acknowledged when interpreting the findings. First, like chlorine-based disinfectants, potassium permanganate is particularly effective against bacteria and viruses but less so against helminth eggs, whose protective shell resists oxidation. Helminth eggs adhering to vegetables must be removed through rubbing during the first wash. Although some studies suggest that potassium permanganate can reduce certain pesticide residues, the removal of pesticides and other contaminants of emerging concern falls outside the scope of this article.

Second, considerations of the broader institutional environment in which this behaviour occurs are also beyond this article’s scope. Factors such as inadequate access to safe water and washing infrastructure, as well as vendors’ limited exposure to clear guidance, shape food safety practices. Similarly, the availability, accessibility, and affordability of potassium permanganate are crucial considerations that are not investigated further here.

Third, vendors were reluctant to allocate significant time to the survey, limiting the inclusion of additional control questions, which may have affected the depth of responses or the ability to cross-validate certain issues. This constraint may have increased the susceptibility to social desirability bias, which refers to the tendency of respondents to answer questions in a manner that will be viewed favourably by others.

Finally, despite extensive preparation work, language and cultural differences posed challenges during survey administration. Conveying the concept of food safety as distinct from food handling practices, and phrasing survey items in a way that matched Likert scale gradations while maintaining clarity, proved challenging. The scale that best captured meaningful distinctions that respondents could make was selected, resulting in the use of 3-, 4-, or 5-point Likert scales across different constructs, favouring practical robustness over theoretical purity. The outcome variable was measured on a 3-point scale (not committed at all, a little committed, very committed), which represents both a pragmatic adaptation to field conditions and a methodological limitation. While ordinal logistic regression successfully modelled this outcome structure, a 5-point or 7-point scale would have provided a more granular measurement and potentially enabled the detection of additional nuanced relationships. The constrained scale reflects the broader challenge in cross-cultural research of balancing statistical ideals with respondent comprehension and cultural appropriateness of measurement instruments.

## Conclusions

This study examined hygiene practices among street food vendors and identified the psychosocial factors that significantly predict the adoption of safer vegetable-washing methods. The primary objective was to determine which psychosocial factors make the use of potassium permanganate more likely than salt for removing faecal pathogens from raw vegetables consumed as salads.

Our results suggest that, while street food vendors value hygiene and recognise the health risks associated with consuming contaminated salads, their understanding of these risks is incomplete and occasionally inaccurate. This leads to misconceptions that hinder the adoption of safer practices. The widespread use of salt for washing vegetables, a practice deeply ingrained in Ghanaian culture, may be the best available option to wash vegetables that will be cooked, but it is inadequate for removing faecal pathogens from vegetables that will be eaten raw. More effective alternatives include washing vegetables under running water or using disinfectants, such as chlorine-based products or potassium permanganate (24,29,31,65). Beyond limited access to piped water, measuring cultural resistance to chlorine-based disinfectants leads us to conclude that it is necessary to explore alternatives, such as potassium permanganate, which is a common vegetable sanitiser in other LMIC.

Although vendors acknowledge the benefits of using a more effective disinfectant than salt, few are familiar with potassium permanganate, and many are cautious about adopting it. Traditional food safety interventions emphasise education, training, and subsidies, but these alone are insufficient to drive widespread adoption. Our findings suggest that personal norms, injunctive norms, and confidence to resume the practice after interruptions are drivers of vendors’ commitment to using potassium permanganate. Public health authorities seeking to promote its use should integrate behaviour change strategies that reinforce personal and injunctive norms, such as leveraging trusted figures of authority and community champions to endorse the practice. This approach is especially relevant in a context like urban Ghana, where social conformity and respect for authority figures strongly shape everyday decisions. Behaviour is guided by what is seen as socially acceptable and endorsed by the approval of trusted community figures, making social norms a powerful lever for change.

These findings are relevant beyond Accra, not only elsewhere in Ghana but also for cities in other LMIC where faecal contamination of vegetables eaten raw is common. By aligning with WHO recommendations that emphasise consideration of socio-cultural constraints, alongside technical and economic measures (16), this article supports a more holistic approach to food safety in informal urban food markets.

## Data Availability

All data produced in the present study are available upon reasonable request to the authors

## List of abbreviations

DALY: Disability-Adjusted Life Years
GHS: Ghanaian Cedi
JHS: Junior High School
KMnO₄: Potassium Permanganate
LMIC: Low- and Middle-Income Countries
ppm: Parts per million
RANAS: Risk, Attitude, Norms, Abilities, Self-regulation (Behaviour change method)
SHS: Senior High School
USD: United States Dollar
WHO: World Health Organization

## Declarations

### Ethics approval and consent to participate

This work conformed with ethical protocols, which were approved by Loughborough University (ref 2022-7727-12403) and IWMI’s Institutional Review Board in Ghana (ref 2023_04).

### Consent for publication

Not applicable.

### Availability of data and materials

All data generated or analysed during this study are included in this published article and its supplementary information files.

### Authors’ contributions

DG conceived and designed the study under the supervision of KG and RS. DG designed the interview guide with input from KG and RS, and the quantitative questionnaire with input from KG, RS and JS. DG collected, interpreted and analysed the data, with confirmatory statistical analysis from JS. DG designed and drafted the manuscript, which KG, RS and JS revised. All authors read the article and approved the final version.

## Acknowledgements

The authors are grateful to Betty Davidson, Fortune Eshun, Wisdom Nartey, Dianah Odoi, Rauf Issah, Tristy Oteng, and Alexandria Rodgers for their dedication during data collection. Special thanks also go to Mabel Kumah and Arnold Ofoli, who served as invaluable research assistants and played a key role in building trust with participants and navigating cultural nuances. We are also sincerely grateful to Dr Ebenezer Amankwaa at the University of Ghana for facilitating these connections and making the collaboration possible. Finally, the authors thank Dr Pay Drechsel, Dr Olufunke Kofie, and Dr Charity Amponsah at the International Water Management Institute for their support throughout this study.

## Footnotes

1 GHS 10 ≈ USD 0.80 in February 2024 when the data were collected.

