## Supplemental tables S1-S10, and figure S1 for "Street-food vendors and food safety: Behavioural predictors of safe vegetable washing in Accra, Ghana"

### Supplementary material

Table S1 Self-reported behavioural frequencies and factual risk knowledge

| Question | n | % |
|---|---|---|
| Do you always find water and soap when you want to wash your hands at work? | | |
| almost never | 9 | 3.6 |
| sometimes | 41 | 16.3 |
| often | 57 | 22.7 |
| almost always | 144 | 57.4 |
| Do you wash your salad vegetables in a basin or under running water (i.e., under the open tap, not in the basin)? | | |
| basin | 235 | 93.6 |
| running water | 16 | 6.4 |
| Do you add salt to the water when you wash your salad vegetables? | | |
| no | 23 | 9.2 |
| yes | 229 | 90.8 |
| Do you add vinegar to the water when you wash your salad vegetables? | | |
| no | 119 | 47.4 |
| yes | 133 | 52.6 |
| Have you ever washed salad vegetables without salt/vinegar because you forgot or it's finished? | | |
| almost never | 138 | 55.0 |
| sometimes | 89 | 35.5 |
| often | 13 | 5.2 |
| almost always | 11 | 4.4 |
| Can germs on salad vegetables come from… ? | | |
| the soil (farm) [a] | 182 | 72.5 |
| the irrigation water (farm) [a] | 191 | 76.1 |
| the dust in the air [a] | 205 | 81.7 |
| touching vegetables with dirty hands [a] | 235 | 93.6 |
| the flies coming on the salad [a] | 232 | 92.4 |
| How often do you think you or other adults can get sick after eating a salad that has germs? | | |
| Almost never | 9 | 3.6 |
| Sometimes | 47 | 18.7 |
| Often | 98 | 39.0 |
| Almost always | 97 | 38.6 |
| How sick do you think you or other adults can be from eating a salad that has germs? | | |
| Not sick at all | 8 | 3.2 |
| A little sick (runny stomach / diarrhoea) | 70 | 27.9 |
| Quite sick (stomach ache) | 72 | 28.7 |
| Much sick (vomiting) | 75 | 29.9 |

| | | |
|---|---|---|
| Very sick (people go to the pharmacy) | 26 | 10.4 |
| Do you think children can get sick after eating a salad that has germs more easily than adults? | | |
| No | 11 | 4.4 |
| Yes | 240 | 95.6 |
| If a child gets sick from eating a salad that has germs, do you think it is more serious than with adults? | | |
| No | 25 | 10.0 |
| Yes | 226 | 90.0 |
| Do you think cholera can be transmitted through eating a salad with germs? | | |
| No | 19 | 7.6 |
| Yes | 207 | 82.5 |
| I do not know | 25 | 10.0 |
| Do you think typhoid fever can be transmitted through eating a salad with germs? | | |
| No | 38 | 15.1 |
| Yes | 129 | 51.4 |
| I do not know | 84 | 33.5 |

Note. *N*=251. [a] Reflects the number and percentage of participants answering ‘yes’ to this question

Table S2 Knowledge of disinfectants and preferences

| Question | n | % |
|---|---|---|
| Do you know about chlorine tablets, like Aquatabs? | | |
| No | 140 | 55.8 |
| Yes | 111 | 44.2 |
| Do you know about potassium permanganate? | | |
| No | 212 | 84.5 |
| Yes | 39 | 15.5 |
| What would be your preferred disinfectant to replace salt? | | |
| Chlorine (Aquatabs tablets) | | |
| 1st choice | 193 | 76.9 |
| 2nd choice | 40 | 15.9 |
| 3rd choice | 1 | 0.4 |
| rejected | 17 | 6.8 |
| Potassium permanganate [a] | | |
| 1st choice | 38 | 15.4 |
| 2nd choice | 164 | 66.4 |
| 3rd choice | 7 | 2.8 |
| rejected | 40 | 16.2 |
| Liquid bleach (Parazone) [b] | | |

| | | |
|---|---|---|
| 1st choice | 1 | 0.4 |
| 2nd choice | 1 | 0.4 |
| 3rd choice | 18 | 7.4 |
| rejected | 224 | 91.8 |

Note. N=251 except [a] N=249, and [b] N=244

Table S3 Attitude, willingness to pay and social norms affecting the use of potassium permanganate

| Question | n | % |
|---|---|---|
| What is most important to you? That your salads are... | | |
| ... plentya | | |
| 1st choice | 9 | 3.7 |
| 2nd choice | 41 | 16.7 |
| 3rd choice | 196 | 79.7 |
| ... safeb | | |
| 1st choice | 183 | 74.1 |
| 2nd choice | 55 | 22.3 |
| 3rd choice | 9 | 3.6 |
| ... tastyb | | |
| 1st choice | 54 | 21.9 |
| 2nd choice | 153 | 61.9 |
| 3rd choice | 40 | 16.2 |
| If tomorrow, you had to start washing salad vegetables with potassium permanganate, how would that make you feel? | | |
| Very worried | 41 | 16.3 |
| A little worried | 55 | 21.9 |
| I'm not sure, a bit worried, a bit happy | 55 | 21.9 |
| A little comfortable | 45 | 17.9 |
| Very comfortable | 55 | 21.9 |
| Washing salad vegetables with potassium permanganate makes salad safer than salt. How advantageous do you think that is? | | |
| Not advantageous at all | 35 | 13.9 |
| A little advantageous | 121 | 48.2 |
| Very advantageous | 95 | 37.8 |
| Do you think using potassium permanganate instead of salt could be a problem? | | |
| Not a problem at all | 92 | 36.7 |
| A small problem | 110 | 43.8 |
| A big problem | 49 | 19.5 |

| | | |
|---|---|---|
| How much of a problem do you think using potassium permanganate could be if potassium permanganate is a bit expensive? | | |
| Not a problem at all | 52 | 20.7 |
| A small problem | 104 | 41.4 |
| A big problem | 95 | 37.8 |
| …if potassium permanganate is hard to find? | | |
| Not a problem at all | 29 | 11.6 |
| A small problem | 84 | 33.5 |
| A big problem | 138 | 55.0 |
| … if your customers or other vendors cause trouble because of it? | | |
| Not a problem at all | 28 | 11.2 |
| A little problem | 68 | 27.1 |
| A big problem | 155 | 61.8 |
| If potassium permanganate is easy to find and your customers like it, how much would you be willing to pay per day to be sure you are washing salad vegetables safely? | | |
| 50 pesewas | 31 | 12.4 |
| 1 cedis | 57 | 22.7 |
| 2.5 cedis | 33 | 13.1 |
| 5 cedis | 61 | 24.3 |
| more than 5 cedis | 69 | 27.5 |
| How many other street food vendors would be willing to use potassium permanganate if it makes salads safer | | |
| No one | 14 | 5.6 |
| A few of them | 165 | 65.7 |
| Many of them | 55 | 21.9 |
| Almost all of them | 17 | 6.8 |
| People who are important to you, like family members, or religious leaders, how much would they approve that you wash your salad vegetables with potassium permanganate? | | |
| They would not approve | 41 | 16.3 |
| I'm not sure | 103 | 41.0 |
| They would approve | 107 | 42.6 |
| To you personally, how important is it to make salad safer by washing vegetables with potassium permanganate? | | |
| Not important at all | 23 | 9.2 |
| A little important | 107 | 42.6 |
| Very important | 121 | 48.2 |

Note. N=251 except [a] N=246, and [b] N=247

Table S4 Abilities and Self-regulation

| Question | n | % |
|---|---|---|
| How confident are you that you would be able to measure the tip of a knife of potassium permanganate? | | |
| Not confident at all | 16 | 6.4 |
| A little confident | 91 | 36.3 |
| Very confident | 144 | 57.4 |
| How confident are you that you would be able to measure 5 litres of water? | | |
| Not confident at all | 19 | 7.6 |
| A little confident | 71 | 28.3 |
| Very confident | 161 | 64.1 |
| How confident are you that you would be able to measure 10 minutes? | | |
| Not confident at all | 20 | 8.0 |
| A little confident | 65 | 25.9 |
| Very confident | 166 | 66.1 |
| How confident are you that you would be able to use potassium permanganate continuously? | | |
| Not confident at all | 49 | 19.5 |
| A little confident | 112 | 44.6 |
| Very confident | 90 | 35.9 |
| If you had to stop using potassium permanganate because it is finished at the pharmacy, how confident are you that you would start using it again? | | |
| Not confident at all | 56 | 22.3 |
| A little confident | 115 | 45.8 |
| Very confident | 80 | 31.9 |
| Could you explain how you would plan to use potassium permanganate? | | |
| No | 54 | 21.5 |
| Yes | 197 | 78.5 |
| How difficult or easy do think it would be to always use the correct quantity of potassium permanganate and water and the correct time when washing salad vegetables? | | |
| Very difficult | 14 | 5.6 |
| A little difficult | 83 | 33.1 |
| Easy | 103 | 41.0 |
| Very easy | 51 | 20.3 |
| If you runout of potassium permanganate and cannot get hold of any in your usual pharmacy, what is your favourite solution? | | |
| go back to using salt/vinegar and give up on using potassium permanganate [a, b] | 143 | 57.4 |
| check at your usual pharmacy until they have potassium permanganate again [a, b] | 32 | 12.9 |

| | | |
|---|---|---|
| check several pharmacies until you find one that has potassium permanganate [a, b] | 10 | 4.0 |
| make sure to buy potassium permanganate at the pharmacy before it is finished at home [a, b] | 64 | 25.7 |
| If these people told you that washing vegetables with potassium permanganate makes salads very safe, how much do you think you would stick to using it? | | |
| Not committed at all | 27 | 10.8 |
| A little committed | 107 | 42.6 |
| Very committed | 117 | 46.6 |

Note. N=251, except [a] N=249. [b] Reflects the number and percentage of participants answering 'yes' to this question

Table S5 Correlations of predictors with outcome

| Predictor | Spearman r | Absolute r | Flag |
|---|---|---|---|
| BL_21_Norm_Personal | .624 | .624 | ** |
| BL_20_Norm_Injunktiv | .565 | .565 | ** |
| BL_23_Maintanance | .532 | .532 | ** |
| BL_15_beliefs_advantage | .519 | .519 | ** |
| BL_24_confidence_recovery | .513 | .513 | ** |
| BL_14_Feelings | .436 | .436 | * |
| BL_19_Norm_Deskriptiv | .382 | .382 | * |
| BL_18_Willing_Pay | .369 | .369 | * |
| BL_16_beliefs_problem | .354 | .354 | * |
| BL_26_action_control | .333 | .333 | * |
| BL_22c_confidence_perf_time | .332 | .332 | * |
| BL_22a_confidence_performance | .301 | .301 | * |
| BL_22b_confidence_perf_volume | .298 | .298 | |
| BL_7_Vulnerability | .272 | .272 | |
| BL_6_Knowledge | .26 | .26 | |
| BL_25_action_plan_1 | .233 | .233 | |
| BL_17a_beliefs_expensive | .215 | .215 | |
| BL_12_beliefs | .103 | .103 | |
| BL_17b_beliefs_hard | -.102 | .102 | |
| BL_17c_beliefs_acceptance | .08 | .08 | |
| BL_8_Severity | -.001 | .001 | |

** Strong association, * Moderate association

Table S6 VIF scores

| Predictor | VIF | Status |
|---|---|---|
| BL_22c_confidence_perf_time | 4.85 | * |
| BL_22b_confidence_perf_volume | 4.53 | * |
| BL_22a_confidence_performance | 2.79 | |
| BL_23_Maintanance | 2.77 | |
| BL_21_Norm_Personal | 2.74 | |
| BL_15_beliefs_advantage | 2.59 | |
| BL_16_beliefs_problem | 2.53 | |
| BL_14_Feelings | 2.42 | |
| BL_26_action_control | 2.31 | |
| BL_20_Norm_Injunktiv | 2.18 | |
| BL_24_confidence_recovery | 2.13 | |
| BL_17a_beliefs_expensive | 1.66 | |
| BL_18_Willing_Pay | 1.56 | |
| BL_19_Norm_Deskriptiv | 1.5 | |
| BL_17b_beliefs_hard | 1.49 | |
| BL_25_action_plan_1 | 1.46 | |
| BL_17c_beliefs_acceptance | 1.45 | |
| BL_6_Knowledge | 1.31 | |
| BL_7_Vulnerability | 1.31 | |
| BL_12_beliefs | 1.2 | |
| BL_8_Severity | 1.14 | |

* Moderate

Table S7 Univariate Kruskal-Wallis tests

| Predictor | Chi Square | df | p value | Significant |
|---|---|---|---|---|
| BL_7_Vulnerability | 20.62 | 2 | < .001 | *** |
| BL_15_beliefs_advantage | 70.68 | 2 | < .001 | *** |
| BL_16_beliefs_problem | 33.34 | 2 | < .001 | *** |
| BL_14_Feelings | 49.69 | 2 | < .001 | *** |
| BL_18_Willing_Pay | 37.88 | 2 | < .001 | *** |
| BL_19_Norm_Deskriptiv | 41.11 | 2 | < .001 | *** |
| BL_20_Norm_Injunktiv | 80.14 | 2 | < .001 | *** |
| BL_21_Norm_Personal | 97.39 | 2 | < .001 | *** |
| BL_22a_confidence_performance | 22.66 | 2 | < .001 | *** |
| BL_22b_confidence_perf_volume | 23.54 | 2 | < .001 | *** |
| BL_22c_confidence_perf_time | 27.82 | 2 | < .001 | *** |
| BL_23_Maintanance | 74.27 | 2 | < .001 | *** |
| BL_24_confidence_recovery | 67.41 | 2 | < .001 | *** |
| BL_26_action_control | 28.49 | 2 | < .001 | *** |

| | | | | |
|---|---|---|---|---|
| BL_25_action_plan_1 | 18.95 | 2 | < .001 | *** |
| BL_6_Knowledge | 17.06 | 2 | < .001 | *** |
| BL_17a_beliefs_expensive | 11.99 | 2 | .0025 | *** |
| BL_12_beliefs | 7.09 | 2 | .0288 | *** |
| BL_17c_beliefs_acceptance | 6.08 | 2 | .0478 | *** |
| BL_17b_beliefs_hard | 2.61 | 2 | .2712 | |
| BL_8_Severity | 0.71 | 2 | .7007 | |

*** p<0.05

Table S8 Comparison of regression methods

| Predictor | Ordinal logistic regression | | | | Linear regression | | | Binary logistic regression Primary coding (1 vs 2-3) | | | | Binary logistic regression Alternative coding (1-2 vs 3) | | | |
|---|---|---|---|---|---|---|---|---|---|---|---|---|---|---|---|
| | β | p | OR | sig | β | p | sig | β | p | OR | sig | β | p | OR | sig |
| BL_21_Norm_Personal | 1.3173 | .0004 | 3.7333 | *** | 0.3104 | .0001 | *** | 1.2397 | .1177 | 3.455 | | 1.4928 | .0005 | 4.4495 | *** |
| BL_24_confidence_recovery | 0.921 | .0021 | 2.5118 | *** | 0.1827 | .0029 | *** | 1.6628 | .0429 | 5.274 | *** | 0.9062 | .0096 | 2.4749 | *** |
| BL_17b_beliefs_hard | -0.5647 | .0378 | 0.5685 | *** | -0.102 | .0582 | | -1.0172 | .0921 | 0.362 | | -0.4633 | .1563 | 0.6292 | |
| BL_20_Norm_Injunktiv | 0.5989 | .0394 | 1.8201 | *** | 0.1177 | .0589 | | -0.1549 | .7947 | 0.856 | | 0.748 | .033 | 2.1128 | *** |
| BL_6_Knowledge | 0.1792 | .0693 | 1.1963 | | 0.0332 | .0994 | | 0.2674 | .1986 | 1.307 | | 0.191 | .0902 | 1.2105 | |
| BL_12_beliefs | 0.5526 | .1401 | 1.7378 | | 0.1214 | .1068 | | 2.0075 | .0167 | 7.445 | *** | 0.2431 | .5814 | 1.2752 | |
| BL_16_beliefs_problem | -0.4846 | .1639 | 0.6159 | | -0.088 | .1844 | | -0.9451 | .3256 | 0.389 | | -0.6815 | .0928 | 0.5059 | |
| BL_18_Willing_Pay | 0.1682 | .2127 | 1.1832 | | 0.0354 | .192 | | 0.4337 | .2078 | 1.543 | | 0.0949 | .5377 | 1.0995 | |
| BL_22c_confidence_perf_time | 0.6408 | .2061 | 1.898 | | 0.1209 | .251 | | 2.101 | .0829 | 8.174 | | 0.2879 | .6211 | 1.3336 | |
| BL_23_Maintanance | 0.3997 | .2199 | 1.4914 | | 0.0783 | .2602 | | 0.8127 | .2892 | 2.254 | | 0.1508 | .6834 | 1.1628 | |
| BL_15_beliefs_advantage | 0.2792 | .4354 | 1.3221 | | 0.0684 | .3416 | | 1.5063 | .1143 | 4.51 | | 0.0805 | .8381 | 1.0838 | |
| BL_19_Norm_Deskriptiv | 0.2875 | .3417 | 1.3331 | | 0.0483 | .3797 | | 1.7847 | .0397 | 5.958 | *** | 0.0829 | .8016 | 1.0864 | |
| BL_26_action_control | 0.1277 | .6499 | 1.1362 | | 0.0301 | .5846 | | 0.084 | .9039 | 1.088 | | 0.1117 | .7304 | 1.1182 | |
| BL_17a_beliefs_expensive | 0.1875 | .4945 | 1.2062 | | 0.0279 | .5945 | | 0.4644 | .4887 | 1.591 | | 0.3704 | .2388 | 1.4483 | |
| BL_7_Vulnerability | -0.0937 | .6644 | 0.9106 | | -0.0198 | .6326 | | -1.0592 | .0798 | 0.347 | | 0.0752 | .7547 | 1.0781 | |
| BL_8_Severity | -0.036 | .8285 | 0.9646 | | -0.0137 | .6571 | | -0.0446 | .9119 | 0.956 | | -0.0517 | .782 | 0.9496 | |
| BL_22a_confidence_performance | -0.244 | .541 | 0.7835 | | -0.036 | .662 | | -0.2749 | .7325 | 0.76 | | -0.0215 | .9654 | 0.9787 | |
| BL_22b_confidence_perf_volume | -0.1489 | .7683 | 0.8617 | | -0.0295 | .7732 | | -1.3397 | .312 | 0.262 | | 0.3568 | .5321 | 1.4288 | |
| BL_25_action_plan_1 | -0.2245 | .6034 | 0.7989 | | -0.0184 | .8365 | | -0.9682 | .2619 | 0.38 | | -0.4175 | .4285 | 0.6587 | |
| BL_17c_beliefs_acceptance | -0.081 | .7651 | 0.9222 | | -0.0073 | .8907 | | 1.1912 | .1771 | 3.291 | | -0.3261 | .288 | 0.7217 | |
| BL_14_Feelings | 0.0424 | .8054 | 1.0433 | | -0.0027 | .9361 | | -0.2638 | .5636 | 0.768 | | 0.0622 | .7476 | 1.0642 | |

*Note.* In the binary logistic regressions, the primary outcome coding was low/no commitment [1-2] vs. high commitment [3]; the alternative outcome coding was not committed [1] vs. any commitment [2-3]; Beta = standardised coefficient; OR = odds ratio (for logistic models only); p = p-value; sig = significant at $p < .05$ (marked with ***). All models include the same 17 predictors with n = 251.

Table S9 Path coefficients for ordinal mediation model

| Path | Coefficient | SE | p value |
|---|---|---|---|
| a1 (Feelings →Personal Norms) | 0.269 | 0.024 | < .001 |
| a2 (Feelings → Injunctive Norms) | 0.249 | 0.029 | < .001 |
| b1 (Personal Norms → Commitment) | 1.554 | 0.307 | < .001 |
| b2 (Injunctive Norms → Commitment) | 0.929 | 0.255 | .0003 |
| c' (Feelings →Commitment, direct) | 0.206 | 0.127 | .1038 |
| c (Feelings → Commitment, total) | 0.712 | 0.104 | < .001 |

*Note.* SE = Standard Error. All path coefficients are in log-odds units appropriate for ordinal logistic regression. $p < .05$ indicates significance. Paths a1 and a2 represent the effect of feelings on each mediator (Personal and Injunctive Norms). Paths b1 and b2 represent the effect of each mediator on commitment, controlling for feelings. Path c' represents the direct effect of feelings on commitment after accounting for mediators. Path c represents the total effect of feelings on commitment without mediators. The non-significant direct effect (c') alongside significant indirect effects indicates complete mediation through normative pathways.

Table S10 Indirect effects with bootstrap confidence intervals (10,000 samples)

| Effect | Estimate | SE | 95% CI | p value |
|---|---|---|---|---|
| Direct effect (c') | 0.206 | 0.127 | NA | .1038 |
| Indirect via Personal Norms (a1*b1) | 0.418 | NA | [0.251, 0.627] | NA |
| Indirect via Injunctive Norms (a2*b2) | 0.232 | NA | [0.104, 0.392] | NA |
| Total indirect effect | 0.650 | NA | [0.478, 0.889] | NA |
| Total effect (c) | 0.712 | 0.104 | NA | < .001 |

Note. SE = Standard Error; CI = Confidence Interval; $p < .05$ indicates significance. Standard errors and p-values are not reported for indirect effects because bootstrap confidence intervals are used to assess significance. An indirect effect is considered significant if its 95% CI does not contain zero. The total effect represents the combined effect of direct and indirect pathways.

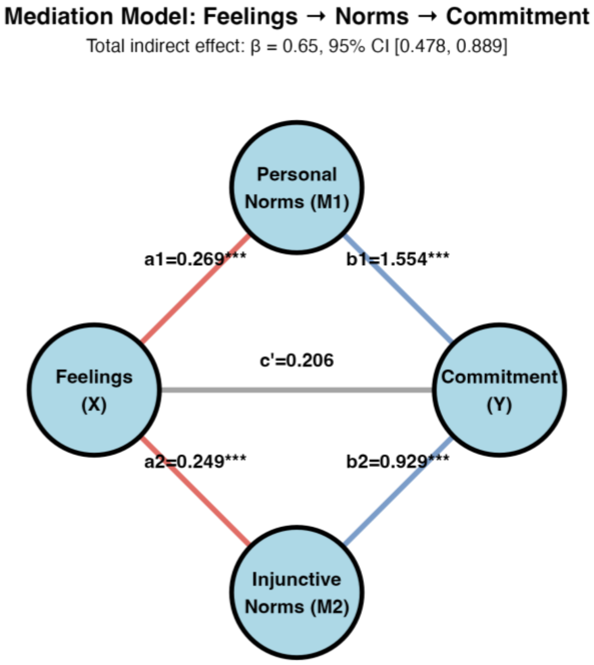


Figure S1 Mediation model diagram
